# Five years functional connectivity reorganization without clinical or cognitive decline in MS

**DOI:** 10.1101/2020.06.19.20135558

**Authors:** Einar August Høgestøl, Samuele Ghezzo, Gro Owren Nygaard, Thomas Espeseth, Piotr Sowa, Mona K. Beyer, Hanne Flinstad Harbo, Lars T. Westlye, Hanneke E. Hulst, Dag Alnæs

**Author notes:** Einar August Høgestøl and Samuele Ghezzo contributed equally to this work as first authors. **Corresponding author:** Einar August Høgestøl, Department of Neurology, Neuroscience Research Unit, Multiple Sclerosis Research Group University of Oslo & Oslo University Hospital, Domus Medica 4, room L-268, Gaustadalleén 34, 0372 Oslo, Norway.

## Abstract

**Objective:** 1) To assess fMRI-based functional connectivity (FC) anomalies in early multiple sclerosis (MS), 2) To determine the relation between FC changes and structural brain damage due to disease progression 3) To study the association between FC changes and cognitive and physical disability.

**Methods:** Structural MRI and resting-state fMRI were acquired from 76 early relapsing-remitting MS patients at baseline (average disease duration 71.7 months ± 63) and after five years. Ninety-four healthy controls (HCs) matched for age and sex were included at baseline. Independent component analysis (ICA) and network modelling were used to measure FC. FC variation was related to expanded disability status scale and neuropsychological outcomes. Brain and lesion volumes were quantified using standard methods. We used the 25 independent components obtained from ICA to estimate the longitudinal stability of the brain connectome as a proxy for functional reorganization over time.

**Results:** The MS subjects were clinically and cognitively stable. Compared to HCs, FC abnormalities were detected within networks and in single connections in patients with early MS at baseline. Over time, FC was relatively invariable, but changes in FC were associated with progression of brain atrophy (ρ = 0.39, p = .06). No significant relationship with clinical and cognitive measures or lesion load was detected.

**Conclusion:** Patients with MS showed evidence of altered FC in the early stages of the disease. Over time, changes in FC seem to be related to a progression of brain atrophy, which are known to precede changes in clinical and cognitive functioning.

## INTRODUCTION

Multiple sclerosis (MS) is a disease characterized radiologically by the accumulation of lesions in grey and white matter over time throughout the CNS^1, 2^. Brain white matter constitutes the structural connectivity backbone supporting large-scale brain functional network connectivity^3, 4^. Accumulating evidence has demonstrated abnormal patterns of brain functional connectivity (FC) in MS patients as compared to healthy controls (HCs)^5-12^. Extensive evidence shows that FC abnormalities are associated with clinical disability in MS^5, 6, 8^-^10, 12^, with a complex pattern of the directionality of FC effects^13^. A longitudinal approach that accounts for all alterations in FC following structural damage in MS is warranted to better understand the complex interplay between disease progression and FC reorganization^13^.

We investigated a prospectively collected MS cohort with comprehensive imaging and clinical data at two time points over five years. First, we explored the baseline resting-state (rs) fMRI MS-data and compared this to a group of age-matched HCs from the same scanner. Then, we used rs-fMRI data to compute the longitudinal stability of each patients’ brain functional connectome^14^, reducing the complex dynamics of the functional connectome into a single, individual-level global marker of longitudinal network reorganization. The resulting rs-fMRI global measure of connectome stability was compared to brain atrophy, lesion load and clinical and cognitive outcomes. Our hypotheses were 1) MS affects FC in early stages of disease, 2) disease progression is associated with FC reorganization over a five-year interval and 3) an individual-level global measure of connectome stability is associated with clinical trajectories in MS.

## METHODS

### Participants

We included 76 MS patients from a prospective longitudinal study at Oslo University Hospital^15-17^. All patients were diagnosed between January 2009 and October 2012 with relapsing-remitting MS (RRMS) according to the revised McDonald Criteria^18^. One patient originally categorized as RRMS was later re-evaluated to be primary-progressive (PP) MS, although with relapses and MRI activity. The patient was excluded from image analysis due to missing rs-fMRI data. Early MS was defined as within the first 10 years of MS symptoms onset. Six participants were not examined with the rs-fMRI sequence at baseline, resulting in 70 patients eligible for the image analyses. All participants were invited for follow-up, five years after the first time point. At follow-up, 62 patients were assessed, with two patients missing the rs-fMRI sequence. Inclusion criteria included age between 18 and 50 years, < 3 years since diagnosis of RRMS at baseline, ≥ 6 weeks since the last relapse or corticoid treatment, fluency in Norwegian language, right handedness, no prior neurological or psychiatric disease, no head injury and no substance abuse. We excluded pregnant or breastfeeding patients and a patient with a previous adverse reaction to gadolinium injection.

Cross-sectional data from HC (n=94, age- and sex-matched with respect to the patient sample) were sampled from the Norwegian Cognitive NeuroGenetics (NCNG) cohort, recruited through newspaper advertisements in the Oslo area^19^. Inclusion criteria were age between 20 and 80 years, no previous diseases affecting the CNS and no previous or current substance abuse.

### Neurological assessment

All MS patients completed a comprehensive neurological examination at baseline and follow-up, including assessment of the expanded disability status scale (EDSS) within the same week as the MRI scan. Physical ability of MS patients was assessed using the Timed 25-Foot Walk (T25-FW) and the 9-hole peg test. No evidence of disease activity (NEDA) -3 was defined as the absence of clinical relapses, new or enlarging MRI lesions and clinical progression as measured by EDSS. First line treatment included interferons, glatiramer acetate, teriflunomide and dimethyl fumarate, while second line treatment included fingolimod, natalizumab and alemtuzumab.

Furthermore, all subjects completed a fatigue self-report questionnaire (FSS)^20^ and underwent cognitive evaluation. FSS mean scores of ≥ 5 were defined as clinically significant fatigue^21^. The cognitive evaluation included tests for processing speed, executive functioning, visuospatial and verbal memory (see Supplemental). In order to obtain a measure of average cognition we first calculated Z-scores for each of the tests administered. Then, we averaged Z-scores within the four cognitive domains previously described, and eventually we averaged the scores across domains, obtaining a measure of overall cognition for each of the MS patients at both time points. Similarly, we averaged the Z-scores for T25-FW and 9-hole peg test to obtain a measure of physical ability.

Cross-sectional California Verbal Learning Test-II (CVLT-II) and colour-word interference test (CWIT) data from the NCNG HC sample and summary statistics for the remaining tests obtained from an Amsterdam HC sample^22^ were used for comparison with the patient group. We considered an annual decline of 0.25 SD in average cognition and physical performance as significant^22^.

### MRI acquisition and structural MRI pre- and post-processing

All MS and HC subjects were scanned using the same 1.5 T scanner (Avanto, Siemens Medical Solutions; Erlangen, Germany) equipped with a 12-channel head coil. For rs-fMRI we used a T2* weighted echo-planar imaging (EPI) sequence (repetition time (TR)=3000 milliseconds (ms), echo time (TE)=70 ms, flip angle (FA)=90°, voxel size=3.44×3.44×4 mm, field-of-view (FOV)=220, descending acquisition, GeneRalized Auto calibrating Partial Acquisition (GRAPPA) acceleration factor=2), 28 transversally oriented slices, no gap, with a scan time of 7 minutes and 30 seconds. The rs-fMRI at baseline included 150 volumes, while the follow-up rs-fMRI scan yielded 100 volumes. The rs-fMRI scan for the HCs constituted of 300 volumes. For fMRI pre-processing and analyses we restricted the rs-fMRI data to the first 100 volumes for all participants. Three dummy volumes were collected to avoid T1 saturation effects. Structural MRI data were collected using a 3D T1-weighted Magnetization Prepared Rapid Gradient Echo (MP-RAGE) sequence (TR=2400 ms, TE=3.61 ms, time to inversion=1000 ms, FA=8°, matrix 192×192, field of view=240, scan time 7 minutes and 42 seconds, sagittal slices=160, voxel size =1.20×1.25×1.25 mm). FLAIR sequence parameters were TR=6000 ms, TE=3.33 ms, time to inversion=2200 ms, FOV=260, FA=variable T2, matrix 256 × 204. Each scan lasted 7 minutes and 2 seconds, sagital slices=176 slices, slice thickness=1 mm, voxel size 1.0×1.0×1.0.

We performed cortical reconstruction by using the T1-weighted scans and volumetric segmentation with FreeSurfer 5.3 (http://surfer.nmr.mgh.harvard.edu/)^23^. To extract reliable volume and thickness estimates, images included in the longitudinal MS MRI dataset were processed with the longitudinal stream in FreeSurfer^24^. To further validate the brain volume measure, we studied its association with lesion-filled masks at baseline (see table e-1 in Supplemental data). Lesion volume was based on LesionQuant output^25, 26^. To validate the lesion output by LesionQuant we additionally assessed its association with lesion masks previously acquired using Cascade and validated by an experienced neuroradiologist at baseline^27^ (see table e-1 in Supplemental data).

### fMRI pre-processing

fMRI was performed using FMRI Expert Analysis Tool (FEAT) Version 6.00, from FMRIB’s Software Library (https://fsl.fmrib.ox.ac.uk/)^28^. Head motion was corrected using MCFLIRT^29^ before linear trends and low-frequency drifts were removed (high-pass filter of 0.01 Hertz). fMRI images were carefully examined by trained research personnel for image artefacts, such as head motion and missing coverage of the scan window. In total, fMRI scans from ten patients and 13 HCs at baseline and eight patients at follow-up were discarded due to incomplete coverage of the visual and parietal cortex. Scans in which the mean relative motion parameter (defined as the average of the translational motion for each scan relative to the previous scan) exceeded 2.5 SD from the average of all runs (across scans) were discarded (none in our MS sample). Brain extraction tool^30^ was used to remove non-brain tissue. Spatial smoothing was performed using a Gaussian kernel filter with a full width at half maximum (FWHM) of 6 mm^31^. FMRIB’s Nonlinear Image Registration tool (FNIRT) was used to register the participants fMRI volumes to Montreal Neurological Institute (MNI) 152 standard using the T1-weighted scan as an intermediate, which had the non-brain tissue removed using procedures for automated volumetric segmentation in Freesurfer 5.3 (http://surfer.nmr.mgh.harvard.edu/)^23^.

Single-session independent component analysis (ICA) was performed for all runs using Multivariate Exploratory Linear Optimized Decomposition into Independent Components (MELODIC)^32^. We used FIX^33^ for automatic classification of the single-session ICA into signal and noise components, in order to regress noise components from fMRI data. Data cleaning also included correction based on the estimated motion parameters for each run, using linear regression. These ICA-based procedures for denoising of fMRI-data efficiently reduce motion induced variability, outperforming methods based on regression of motion parameters, scrubbing or de-spiking^34^.

### ICA and functional connectivity matrices

The cleaned and MNI-conformed rs-fMRI datasets were submitted to temporal concatenation group independent component analysis (gICA) using MELODIC^32^ with a model order of 40. These group level spatial components were then used as spatial repressors against the original rs-fMRI datasets to estimate subject-specific components and associated time series (dual regression)^35^. After removing 15 ICs that originated in non-CNS areas and therefore were classified as noise components by a careful evaluation, we extracted a total of 25 ICs for further analysis. The times series of the noise-ICs were regressed out of the time series of the kept ICs. We then calculated connectivity matrices using full as well as regularized partial correlations with automatic estimation of regularization parameters at the individual level^36, 37^. Based on the Euclidean distances of the full temporal correlations, the ICs grouped into four clusters largely representing (1) and (2) default mode network (DMN) and frontoparietal areas, (3) auditory network and (4) sensory/motor areas (figure 1). For detailed information regarding the specific ICs or the anatomical constitution of the networks, please refer to table e-2 in Supplemental Data. Finally, since partial correlations are assumed to represent direct connections between nodes, these were used in further analyses of case-control differences and connectome stability.

**Figure 1.**
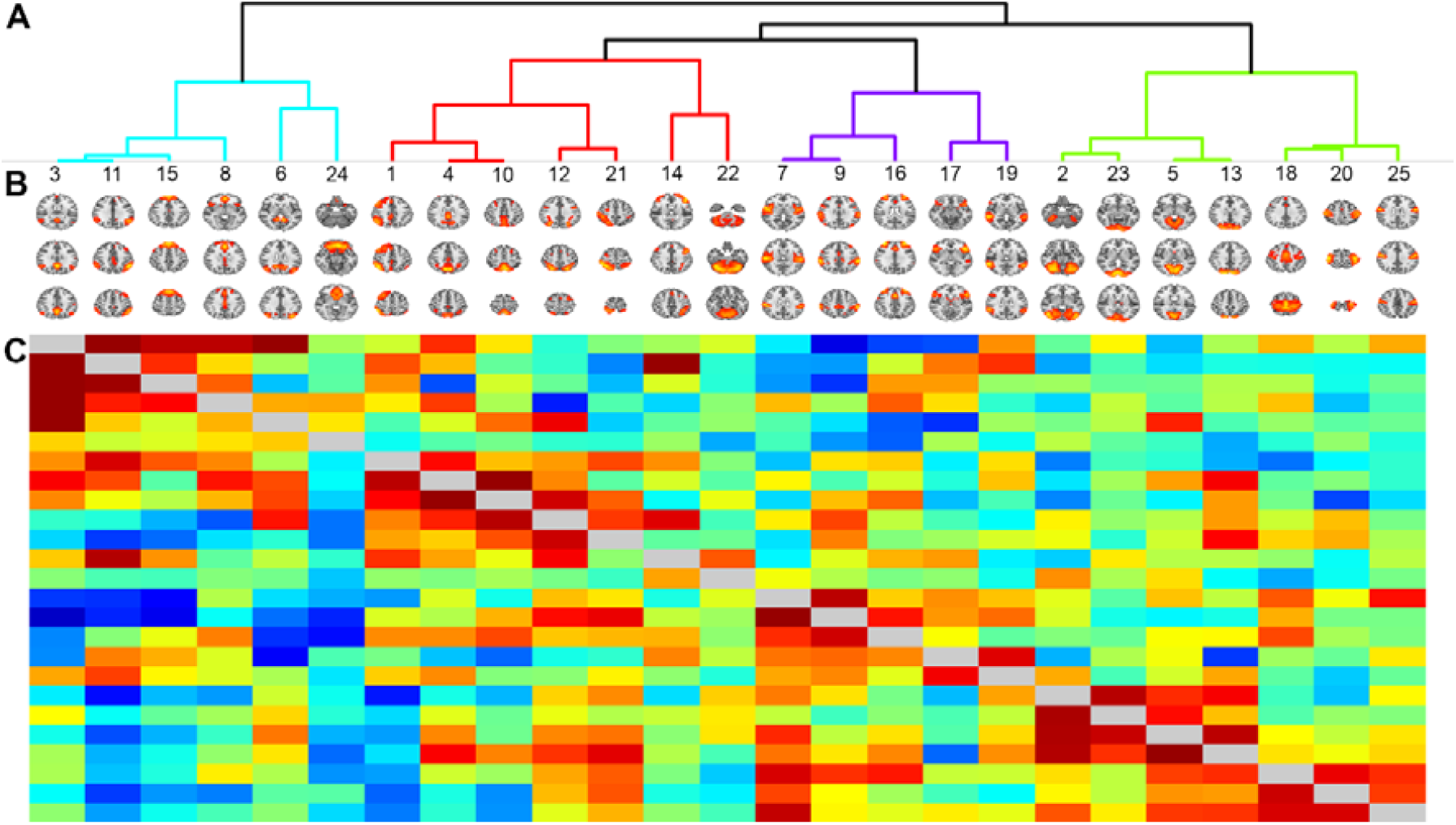
Hierarchical clustering of brain nodes. (A) Dendrogram showing the clustering of the nodes, based on the full correlations across conditions and participants. The colours of the dendrograms represent the four ICA clusters (1), (2), (3) and (4), from left to right. (B) The 25 ICs from group ICA used in the analyses. (C) The full correlation matrix across conditions and participants is shown below the diagonal. Regularized partial correlation across conditions are presented above the diagonal. Warm colours indicate positive correlations, cold colours negative correlations.

### Connectome stability

We computed the stability of the brain functional connectome for each MS patient as a proxy for FC reorganization. We vectorized each participant’s whole-brain connectivity matrix, creating a vector of size 1 × 300 (25 ICs and 300 unique links between them). Then, we calculated the within subjects Spearman correlation coefficient between fMRI at baseline and follow-up, in this way obtaining a measure of whole-brain connectome stability^14^. Similarly, we computed the within-network connectome stability for each of the four clusters previously identified. Alterations in the rank of edges across the connectome between baseline and follow-up, independent of their direction, will be reflected in the connectome stability, providing an individual global and network-specific measure of FC reorganization for the MS patients. To further validate the implementation of the stability of the brain functional connectome as a measure that accounts for all changes in FC over time, we correlated connectome stability with the sum of the squared differences in FC between baseline and FU for all edges in each of the MS patients.

### Statistical analysis

For statistical analyses, we used R^38^ and MATLAB version 2014a (The MathWorks Inc., Natick, MA, 2018). The changes in performance for all cognitive and clinical tests between baseline and follow-up were measured by paired sample t-tests.

We used separate multivariate linear models to test for differences in whole brain FC, within network FC, and FC at the level of single edges between HCs and MS patients at baseline. We performed paired sample t-tests to assess changes in FC over time in the MS cohort.

To investigate whether disease progression was associated with functional reorganization, we used separate multivariate linear models to compare whole brain, and within network, connectome stability between patients that showed evidence of disease activity (EDA) and patients with NEDA. To further test the hypothesis that structural damage due to disease progression is associated with a reduction in the stability of the functional connectome we correlated whole brain functional connectome stability of EDA patients with measures of brain volume loss and accumulation of lesion load over five years. For lesion volumes we used log transformation, plus a constant k = 2, to account for the lack of normal distribution in the resulting volumes.

Finally, we used multivariate linear models to assess the relationship between connectome stability with cognition and physical ability at follow-up, and on the change in these tests between baseline and follow-up.

We adjusted for sex, age, mean relative motion and signal to noise ratio in all linear models. Significance was defined as p < .05 in all the performed tests. In the first part of the study, when we compared FC between HCs and MS patients and later investigated changes in FC over time in the MS cohort, we corrected p value for multiple testing by false discovery rate. Due to the limited sample size of the subgroups, we used permutation testing to correct p value for multiple testing in the comparisons of connectome stability between EDA and NEDA patients. Specifically, we obtained an empirical null-distribution of estimates for the group difference across 1000 permutations randomly permuting the group-label. The family wise error was controlled by collecting the maximum test statistic across the whole brain and within-network tests for each permutation^39^. The resulting p value was calculated by dividing the number of permuted beta-values equal or larger than the point estimate by the total number of permutations.

### Standard protocols approval, registrations, and patient consents

The project was approved by the regional ethical committee of South Eastern Norway (REC ID: 2011/1846, 2016/102 and 2009/2070), and all participants received oral and written information and gave their written informed consent.

### Data availability

Summary data as published in this paper will be available, but other data are not publicly available because of patient privacy restrictions decided by the Regional Ethical Committee. We may apply for permission to share data with new collaborators, still adhering to patient privacy requirements of the “Law of Health Research”. All code needed to replicate our described analyses is available upon request from the corresponding author.

## RESULTS

### Sample characteristics at baseline and follow-up

Our MS sample included 71% (N=54) females, in line with the prevalence of MS in the general population. At follow-up 44% (N=27) of the patients met criteria for NEDA-3. The median EDSS (2.0) score did not change after five years. Only one patient developed secondary progressive MS (SPMS) at follow-up. Mean time between baseline and follow-up was 4.5 years (SD = 0.4 years, range = 3.7-5.4 years). Disease modifying treatment (DMT) were used by 78% and 69% of the patients at baseline and follow-up, respectively (table 1). The proportion of patients using second line treatment increased from 13% at baseline to 32% at follow-up. HCs were matched to the MS sample at baseline by age (HC mean years = 34.89, SD = 9.17) and sex (74% of HC were female). 70% of HCs from the Amsterdam sample were females, (mean age years = 41.86, SD = 11.44).

**Table 1.**
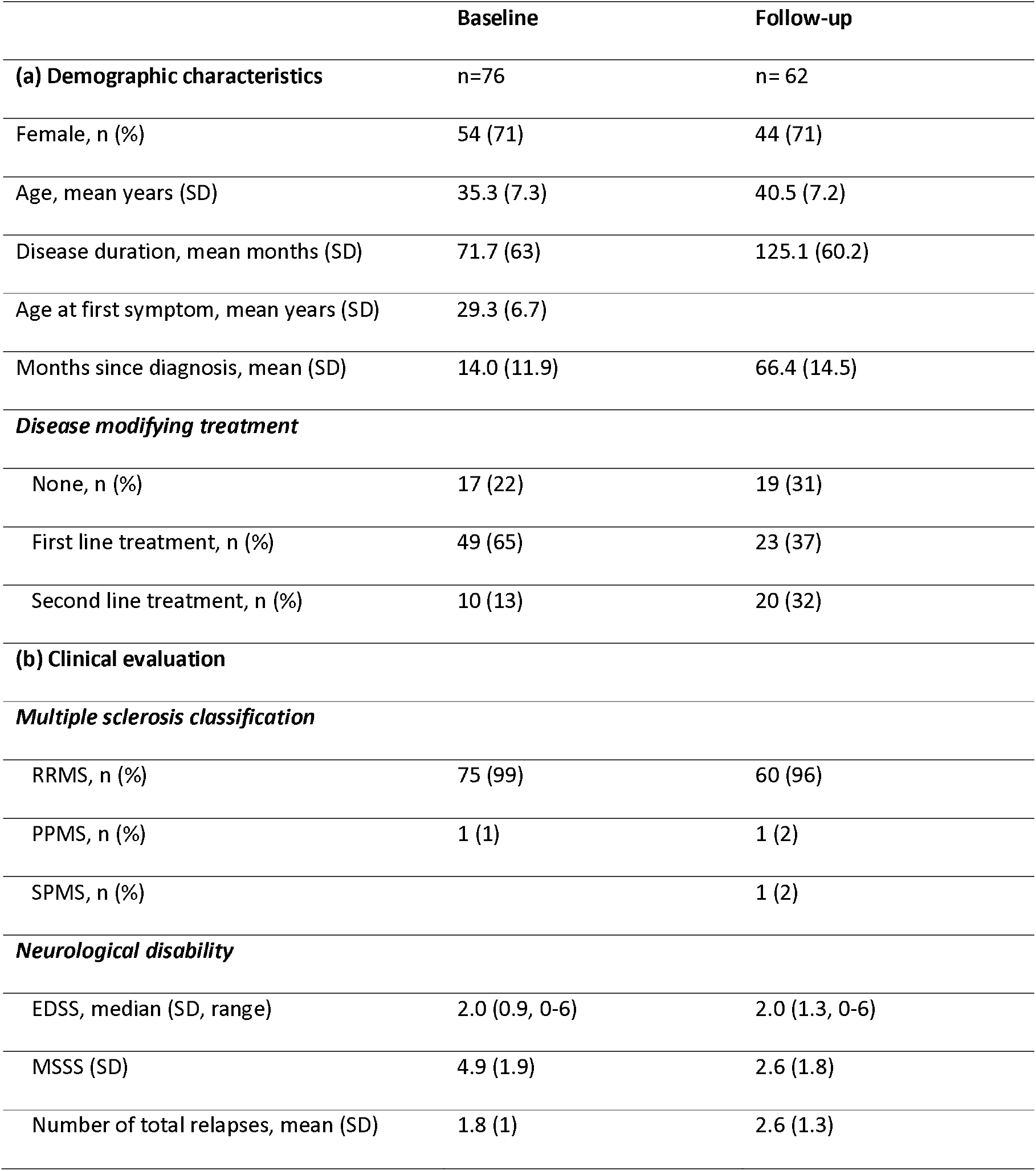
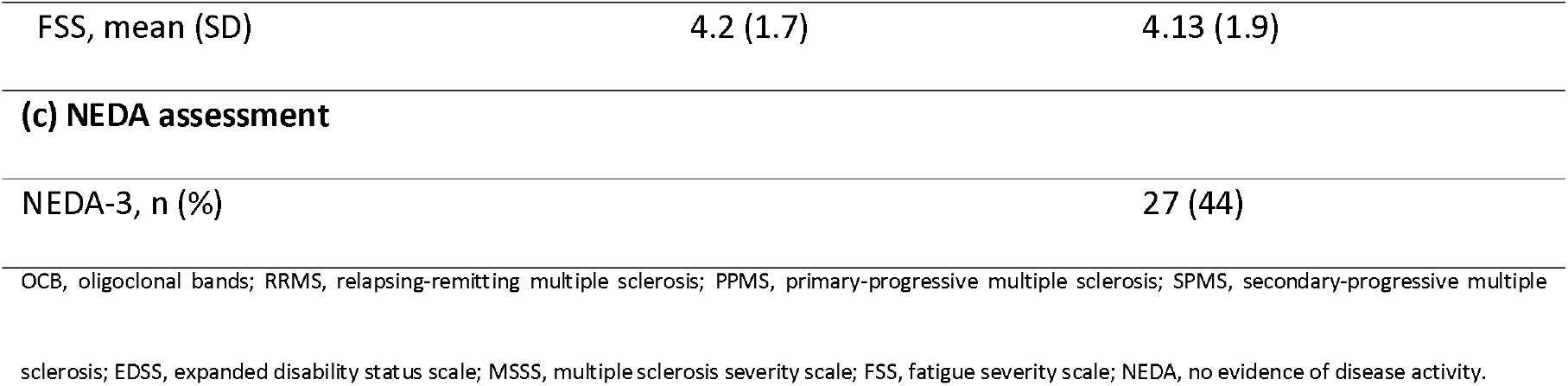
Demographic and clinical characteristics of the multiple sclerosis patients

The performance of MS patients on all cognitive tests of the CWIT, in addition to the CVLT-II, improved over time (table e-3 in Supplemental Data). The performance in symbol digit modalities test, that also was used to investigate processing speed, did not change over time (table e-3 in Supplemental Data). Both T25-FT and 9-hole peg test at follow-up did not show any significant development when compared to baseline (table e-3 in Supplemental Data).

We found no effects of motion parameters such as signal to noise ratio and mean relative motion with clinical and cognitive outcomes (table e-4 in Supplemental Data).

### Functional connectivity abnormalities in MS versus HCs at baseline

Overall FC, for the complete combined functional connectome, was lower in MS patients compared to HCs. FC within three of the four large-scale networks investigated was significantly decreased in MS patients. After correction for multiple testing only network 1 and 2 remained significantly different from controls (table 2). Edge wise analysis showed that a connection (IC11-IC15) in network 1 was weaker in MS relative to controls (β = -0.1, t(135)= -5.21, p = .0002), while another (IC6-IC11) one was stronger in patients compared to HCs (β = 0.07, t(135)= 3.54, p = .032). An edge (IC10-IC14) in network 2 was increased in MS (β = 0.08, t(135)= 3.74, p = .002) and within network 3 one connection (IC7-IC16) decreased intensity in patients relative to controls (β = - 0.08, t(135)= - 3.94, p = .001). Also, a connection (IC2-IC7) between networks 3 and 4 was altered in MS patients (β = - 0.08, t(135)= - 4.02, p = .001). Figure 2 lists the comparisons in FC between MS and HCs at the level of single connections, showing that FC abnormalities are bidirectional in MS (figure e-1 in Supplemental data for complete p values).

**Table 2.**
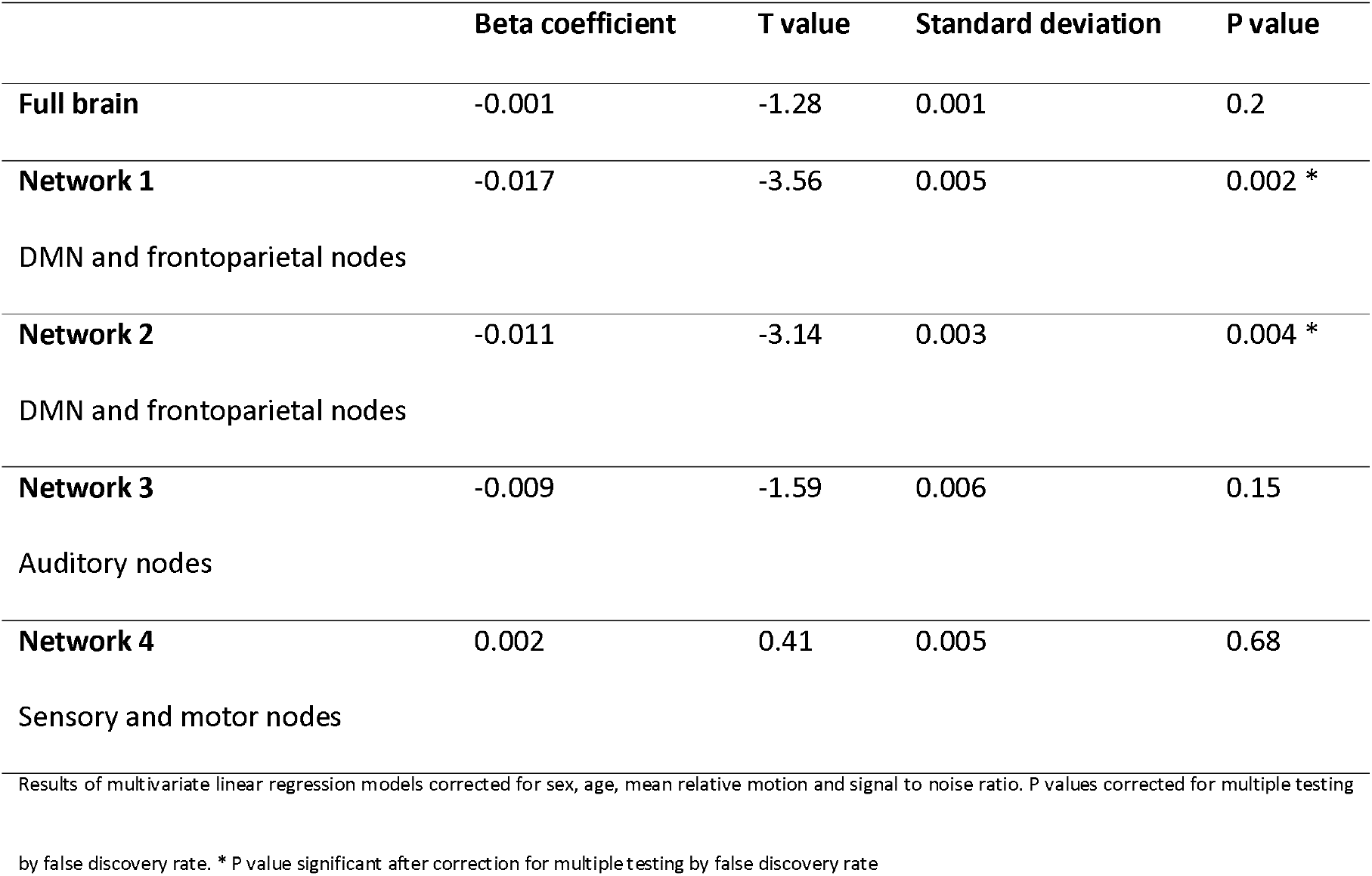
Within network functional connectivity abnormalities in MS

**Figure 2.**
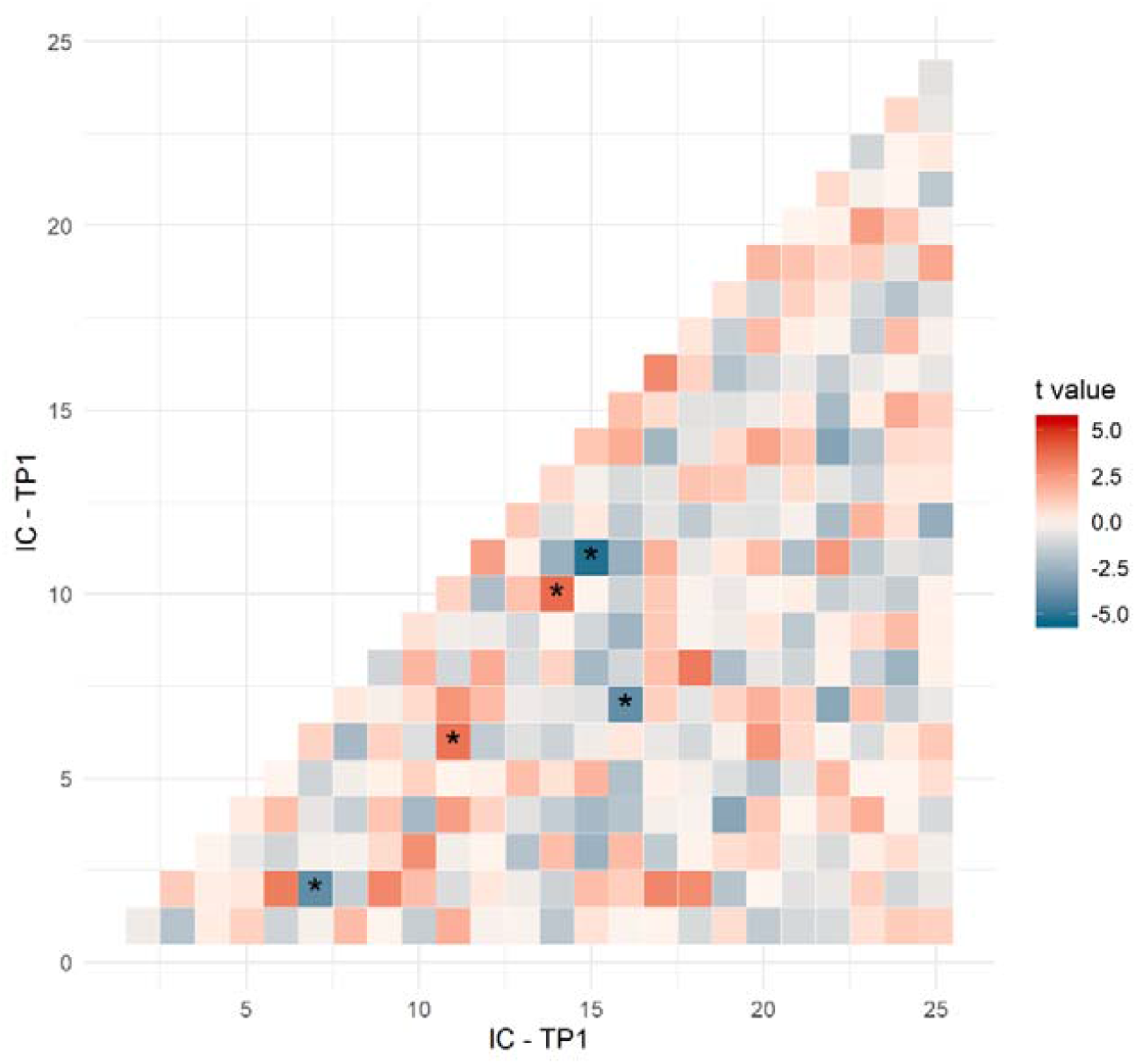
Edgewise analysis of functional connectivity abnormalities in MS vs HCs. T values form multivariate linear regressions assessing differences in FC at the level of single connections between MS and HCs. Warm colours indicate increased FC in MS, cold colours a decrease in FC. * P value significant after correction for multiple testing by false discovery rate.

FC did not change significantly over time in the MS cohort, as measured by paired sample t-test between all ICs considered singularly, and between ICs grouped into networks, across time (figure e-2, e-3 and table e-5 in Supplemental Data). Additional analysis including careful lesion masking during the estimation of the node time series did not change the main effects or interpretation of the results (figure e-4 in Supplemental Data).

### Connectome stability over time

The stability of the brain functional connectome in the whole MS cohort, and in the EDA and NEDA subgroups are depicted in Figure 3, enabling visualization of FC reorganization. Connectome stability of network 2 was lower in EDA patients compared to NEDA patients (β = 0.14, t(34) = 2.26, p = .03), but did not survive correction for multiple testing (table e-6 in Supplemental Data).

**Figure 3.**
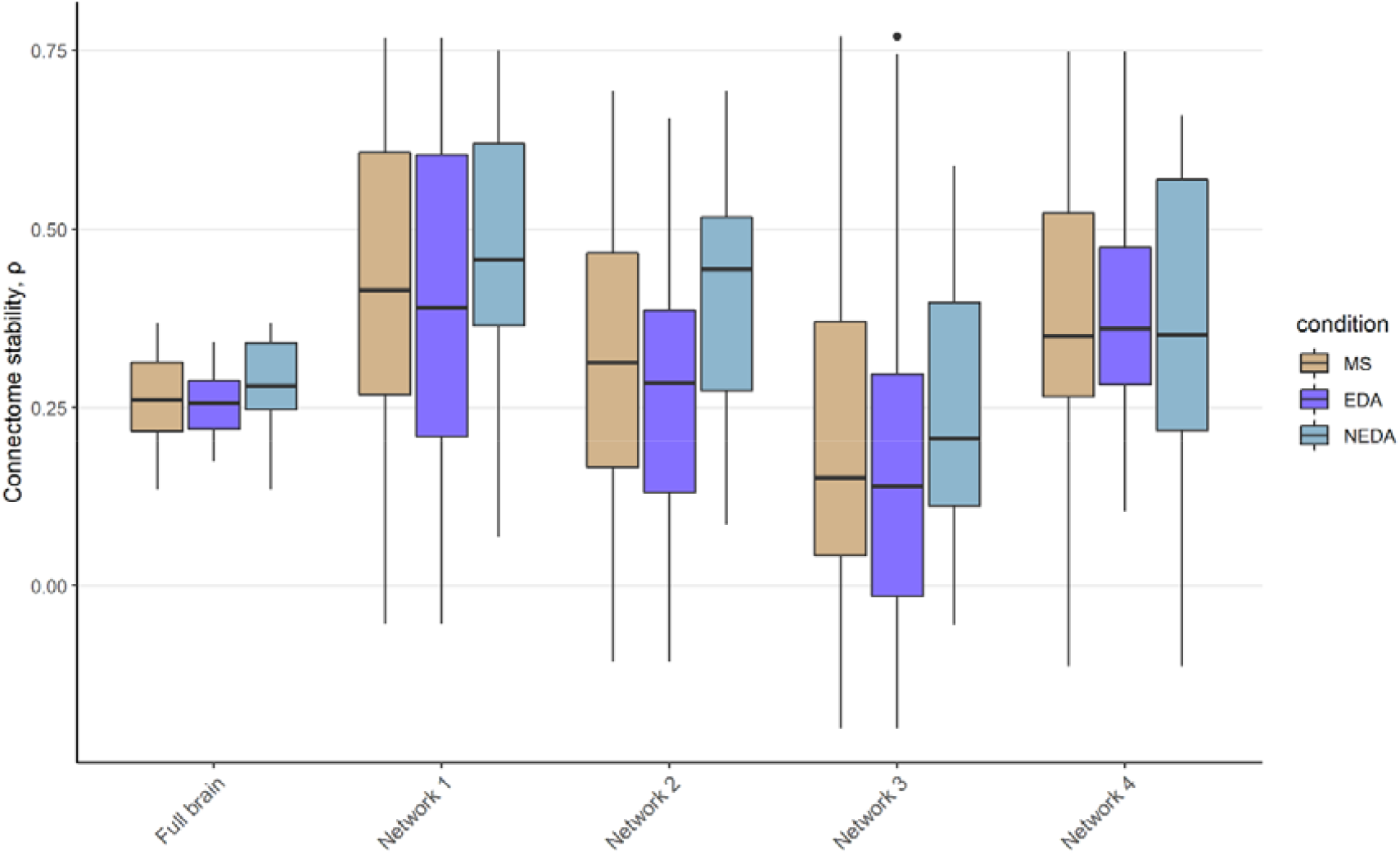
Stability of the brain functional connectome between baseline and follow-up for the MS sample as a whole, and for the subgroups of EDA and NEDA, respectively for the global estimate and all resulting networks. We found a trend were the patients in the EDA subgroup compared to both patients with NEDA and HC patients had improved connectome stability.

The analyses revealed a borderline significant association between connectome stability and change in brain volume between baseline and follow-up, indicating that the larger changes in FC were associated with increased brain volume loss over time, but no significant association was detected with the accumulation of lesion volume over time (figure 4). We also checked for apparent relations between motion parameters such as signal to noise ratio and mean relative motion and brain atrophy and accumulation of lesion load, but we did not find any (table e-7 in Supplemental Data).

**Figure 4.**
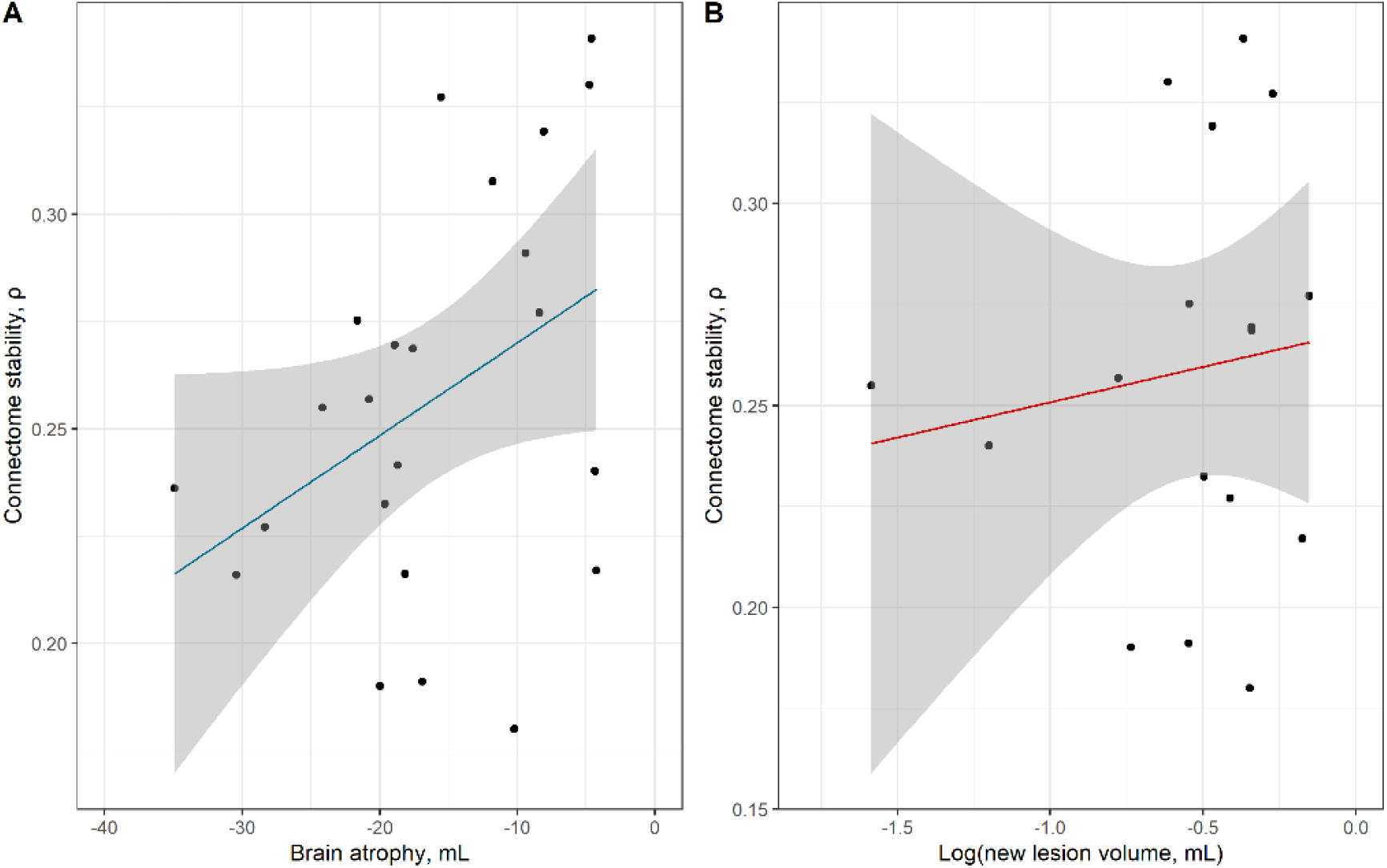
Correlation between structural damage and stability of the brain functional connectome. (A) correlation between brain atrophy and the rate of FC reorganization, ρ = 0.40, p = .06. (B) correlation between accumulation of lesion volume over time and the rate of FC reorganization, ρ = 0.13, p = .63.

We correlated the stability of the brain functional connectome with the sum of the squared differences in FC between baseline and FU, and we found that these two indices were highly related (ρ = - 0.59, p < .0001).

### Clinical relevance of functional connectivity changes

Finally, we explored the effects of the changes in FC on cognitive performance and physical ability using separate general linear models with average cognition and physical ability at FU as dependent variables, covarying for age, sex, signal to noise ratio and mean relative motion. Younger age at FU (β = -0.03, t(34) = -2.08, p = .045) was associated with better cognitive performance at FU (figure 5A). Neither stability of the brain functional connectome, age, sex, signal to noise ratio nor mean relative motion were associated with longitudinal changes in cognitive performance (p > .05) (figure 5B).

**Figure 5.**
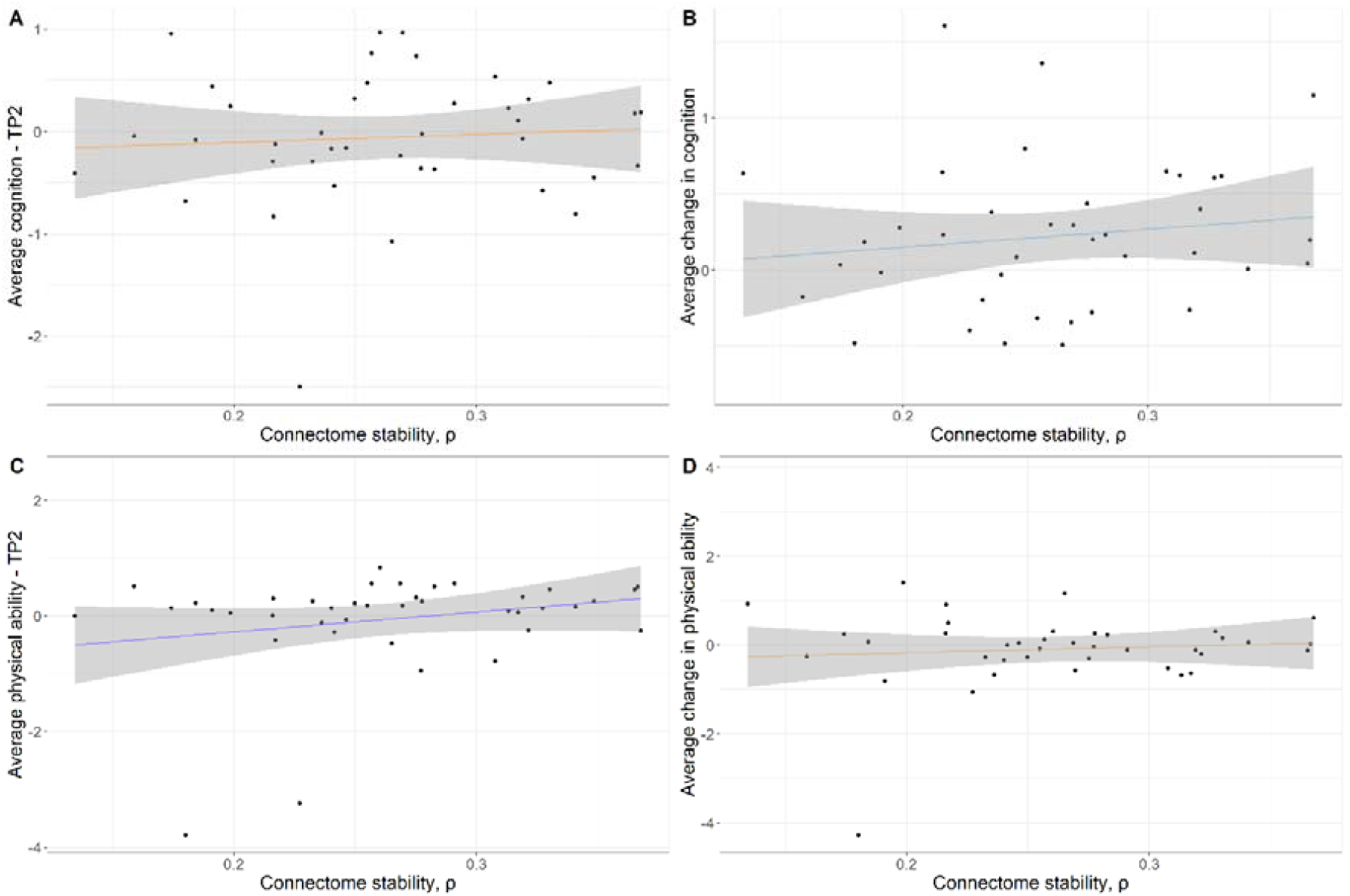
Effect of FC reorganization on cognitive performance and physical ability. (A) Effect of FC reorganization on average cognition at follow-up, β = 1.98, t(34) = 1.20, p = .27. (B) Effect of FC reorganization on average change in cognition, β = 1.00, t(33) = 0.64, p = .52. (C) Effect of FC reorganization on physical ability at follow-up, β = 4.56, t(34) = 2.00, p = .05. (D) Effect of FC reorganization on average change in physical ability, β = 0.89, t(33) = 0.33, p = .75.

For physical ability, lower connectome stability (β = 4.56, t(34) = 2.00, p = .05), higher age at FU (β = -0.05, t(34) = - 2.45, p = .02) and sex, with women scoring better than men, (β = 0.70, t(34) = 2.46, p = .02) were associated with decreased physical ability at FU (figure 5C). None of the investigated variables were associated with changes in physical performance over time (p> .05) (figure 5D).

## DISCUSSION

In this study, we investigated clinical relevance of connectome stability as an individual-level global marker of longitudinal FC changes, in a five years longitudinal prospective study of patients recently diagnosed with MS. In addition, we performed a cross-sectional case-control comparison with a matched group of healthy controls, assessing network- and edgewise FC differences between MS patients and controls.

The case-control comparison replicated previous reports of bi-directional FC differences in MS compared to a matched HC sample^11, 40^, supporting our first hypothesis that FC aberrations are present already in the early stages of MS.

Schoonheim et al. (2010) have proposed a model for functional reorganization of the brain in relation to structural damage and clinical impairment in MS^41^, postulating that, at least in the early stages of MS, functional connectivity would increase as a mechanism to suspend the effects of microstructural damage occurring in the brain as a compensatory mechanism to delay clinical disability. Since then, accumulating evidence has established that FC aberrations are in fact bidirectional in MS^11^, and the authors recognized the first model postulated was overly simplified^13^. Although their intuition and conceptualization of the model for FC reorganization following disease progression in MS was valid, we hypothesize that their measure for FC reorganization was incomplete.

In this study, we present a novel approach enabling us to study individual longitudinal FC reorganization in relation to disease progression in MS. Investigating the longitudinal stability of the brain functional connectome allows us to study the complex dynamics of FC reorganization in MS, finally enabling us to test the model presented by Schoonheim et al.^41^, considering not only the increase in FC, but the whole complex interplay of FC changes following structural damage in MS.

In accordance with our hypothesis that disease progression in MS was associated with FC reorganization as measured by connectome stability, we found that FC changed at a higher rate as brain atrophy, but not lesion volume, increased. In line with the same hypothesis, we expected to see increased FC reorganization in patients experiencing disease activity between baseline and follow-up. Connectome stability in network (2) was lower in EDA patients compared to NEDA patients, however the observed difference between these two sub-groups did not remain significant after correcting for multiple testing.

Finally, we assessed the clinical relevance of the connectome stability measure in our sample. We did not observe significant associations between connectome stability and progression of cognitive and physical impairment. A possible reason might be that our MS cohort was remarkably stable in the follow-up period, exemplified by observations that only two participants displayed a significant decrease (defined as an annual reduction of > 0.25 SD in the five years follow-up period) in physical ability, and none in average cognition (figure e-5 and e-6 in Supplemental data respectively). Lower connectome stability was associated with lower physical ability at follow-up, but the result was likely to be driven by two outliers performing very poorly at follow-up. We did not detect any association between FC reorganization and average cognition at follow-up. According to the model presented by Schoonheim et al^41^, FC reorganization delays the onset of clinical impairment. Since we did not detect any worsening in the clinical performance of our MS patients during the observed period of time, it is tempting to speculate whether the FC changes, that occurred, delayed the onset of impairment. Therefore, it will be important to repeat these analyses in a planned follow-up of the group of patients in a few years.

Using connectome stability as an individual-level longitudinal measure, by incorporating the global dynamics of FC changes in the brain, has previously been used to study mental health in adolescent neurodevelopment^42^ as well as severe mental disorders^14^, but has so far remained unexplored in MS. The measure is based on longitudinal within-subjects Spearman correlation coefficients^14, 42^. Any change in FC that occurs over time, independent of the direction, will cause a shift in ranks that lowers the Spearman correlation coefficient, thus providing a global measure sensitive to relative FC reorganization. However, the shift in ranks of one connection causes the following shift in ranks of all the others, potentially inflating the measure of connectome stability. We correlated the index of connectome stability with the sum of the squared differences in FC to control for possible artefacts due to the implementation of a rank-based measure. Since the correlation is strong, highly significant and goes in the expected direction we are confident that connectome stability is a valid approach to investigate FC reorganization in MS.

Our study is, to our knowledge, one of very few published studies investigating longitudinal FC in MS^40, 43^. In general, our results reproduced and confirmed the results reported by Rocca et al and Koubiyr et al. (2019) investigating clinically isolated syndrome (CIS) patients in a longitudinal setting^40, 43^. All these studies show FC abnormalities at the early phases of the disease and, in line with Rocca et al. we found that FC alterations do not correlate with lesion volume. Furthermore, even though the main approach to study FC reorganization is different in these studies, all data-sets suggest brain reorganization during disease progression.

A limitation of our study is that we could not investigate the difference in connectome stability between HCs and MS patients due to the lack of MRI at follow-up for HCs. Since HCs performed cognitive tests only at baseline, we used the results of HCs at baseline to create the Z-scores for MS patients at follow-up. Z-scores for physical ability were based on analyses of performance of MS patients only. Practice effects in cognitive tests seem to have occurred, but we could not account for them, for example calculating the reliable change index as done by Eijlers et al.^22^. The sample size of our MS cohort is comparable with that of previous studies investigating FC longitudinally^40^. In sub-group analyses the sample sizes are modest. Finally, functional connections of the brain are known to be dynamic, and future studies with longer scan duration may be able to pursue approaches for assessing the dynamic aspects of the FC in the brains of patients with MS.

To conclude, in this longitudinal study we found that our MS cohort was remarkably clinically stable. Only two patients showed a significant decrease in physical ability and cognitive performance was preserved in all participants in the follow-up period. We found that FC abnormalities are bidirectional and occur already at the early stages of MS. We used the stability of the brain functional connectome as a proxy for FC reorganization, enabling us to study FC changes in relation to disease progression. We found that FC reorganization and brain atrophy advance in parallel. In future studies, the MS cohort needs to be followed up again to enable further analyses of functional brain changes and clinical course. Connectome stability enables fMRI data to be condensed into a proxy as an individual marker of brain health in a personalized medical approach to complement the existing biomarkers in MS^44^.

## Data Availability

Summary data as published in this paper will be available, but other data are not publicly available because of patient privacy restrictions decided by the Regional Ethical Committee. We may apply for permission to share data with new collaborators, still adhering to patient privacy requirements of the Law of Health Research. All code needed to replicate our described analyses is available upon request from the corresponding author.

## Acknowledgement

We thank all patients participating in our studies. We acknowledge the collaboration with members of the Multiple Sclerosis Research Group and NORMENT at the University of Oslo and Oslo University hospital, especially Elisabeth G. Celius for her guidance during the investigations of the MS patients and Tobias Kaufmann for valuable help and input on the statistical approach. We also thank the research assistants Kristin Liltved Grønsberg, May-Britt Gjengstø Utheim, Julia Timofeeva, Hedda Maurud, Siren Tønnessen and Petter Espeseth Emhjellen who all contributed in the cognitive testing of the patients.

## Study funding

The project was supported by grants from the South-Eastern Health Authorities of Norway (grant number 2011059/ES563338/Biotek 2021) and by the European Union under the Horizon 2020 programme (grant number 733161).

## Disclosures

E. A. Høgestøl has received honoraria for lecturing from Biogen, Merck and Sanofi-Genzyme, and unrestricted research support from Merck and Sanofi-Genzyme. P. Sowa has received honoraria for lecturing and travel support from Merck. M. K. Beyer has received honoraria for lecturing from Novartis and Biogen Idec. H.F Harbo has received travel support, honoraria for advice or lecturing from Biogen Idec, Sanofi-Genzyme, Merck, Novartis, Roche, and Teva and an unrestricted research grant from Novartis. Hanneke Hulst received honoraria for speaking at scientific meetings, serving at scientific advisory boards and consulting activities from Biogen, Celgene, Genzyme, Merck and Roche. She has received research support from the Dutch MS Research foundation and serves on the editorial board of Multiple Sclerosis Journal (MSJ). S. Ghezzo, G.O. Nygaard, T. Espeseth, L.T. Westlye and D. Alnæs report no disclosures.

## Appendix

### Authors

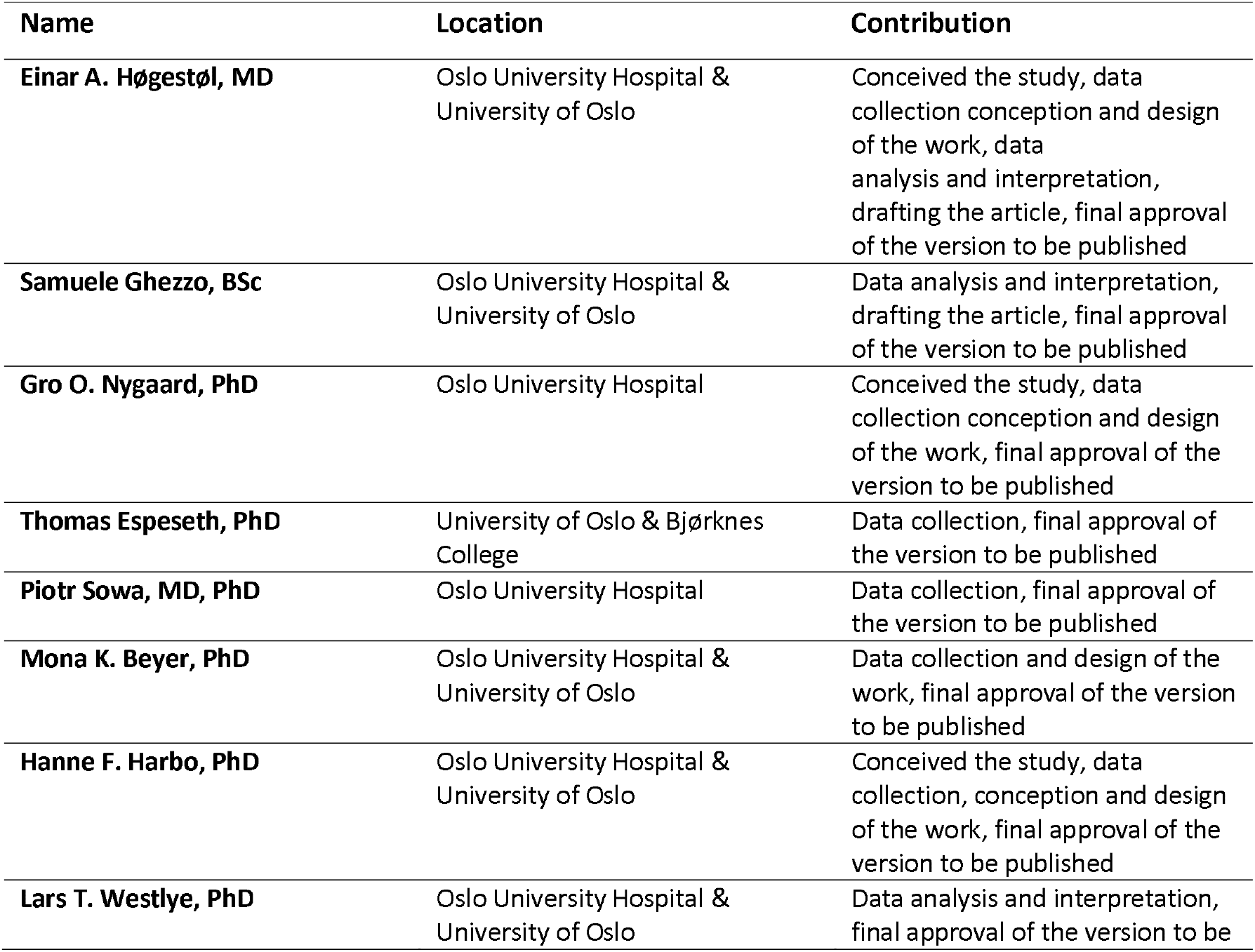
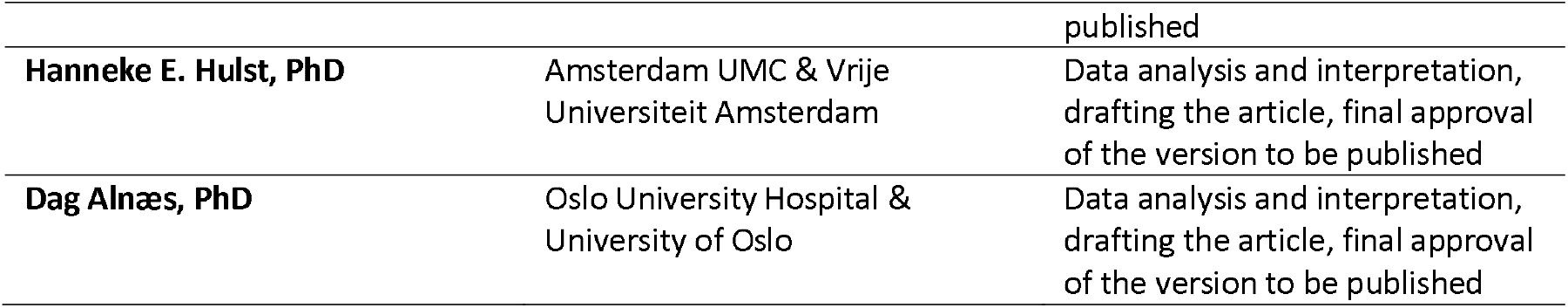

